# Large Language Models Diagnose Facial Deformity

**DOI:** 10.1101/2024.07.11.24310274

**Authors:** Jungwook Lee, Xuanang Xu, Daeseung Kim, Hannah H. Deng, Tianshu Kuang, Nathan Lampen, Xi Fang, Jaime Gateno, Pingkun Yan

## Abstract

**Purpose:** This study examines the application of Large Language Models (LLMs) in diagnosing jaw deformities, aiming to overcome the limitations of various diagnostic methods by harnessing the advanced capabilities of LLMs for enhanced data interpretation. The goal is to provide tools that simplify complex data analysis and make diagnostic processes more accessible and intuitive for clinical practitioners.

**Methods:** An experiment involving patients with jaw deformities was conducted, where cephalometric measurements (SNB Angle, Facial Angle, Mandibular Unit Length) were converted into text for LLM analysis. Multiple LLMs, including LLAMA-2 variants, GPT models, and the Gemini-Pro model, were evaluated against various methods (Threshold-based, Machine Learning Models) using balanced accuracy and F1-score.

**Results:** Our research demonstrates that larger LLMs efficiently adapt to diagnostic tasks, showing rapid performance saturation with minimal training examples and reducing ambiguous classification, which highlights their robust in-context learning abilities. The conversion of complex cephalometric measurements into intuitive text formats not only broadens the accessibility of the information but also enhances the interpretability, providing clinicians with clear and actionable insights.

**Conclusion:** Integrating LLMs into the diagnosis of jaw deformities marks a significant advancement in making diagnostic processes more accessible and reducing reliance on specialized training. These models serve as valuable auxiliary tools, offering clear, understandable outputs that facilitate easier decision-making for clinicians, particularly those with less experience or in settings with limited access to specialized expertise. Future refinements and adaptations to include more comprehensive and medically specific datasets are expected to enhance the precision and utility of LLMs, potentially transforming the landscape of medical diagnostics.

## 1. Introduction

Cephalometric analysis is a fundamental diagnostic tool in orthodontics, oral and maxillofacial surgery [1,2], and craniofacial growth evaluation. It involves the measurement of skeletal, dental, and soft tissue structures, with a focus on assessing facial anatomy through the lengths, ratios, and angles between cephalometric landmarks. Clinicians utilize cephalometric analysis to determine the severity and type of jaw deformities, such as maxillary protrusion or mandibular retrognathia. This analysis plays a crucial role in the planning and executing surgical interventions and treatment strategies [3,4].

Numerous studies have used “norms” of what is considered clinically normal facial morphology to assess facial structure [5,6]. However, these norms are often derived from limited population samples and may not accurately represent the diverse demographics, leading to potential misdiagnoses or misinterpretations [7,8]. Therefore, establishing universally accepted cephalometric standards is challenging due to disagreement among experts on norm interpretation and application, resulting in a lack of uniform, objective criteria for diagnosing jaw deformities [9,10]. The inherent subjectivity in the interpretation of measurements against variable norms can significantly impact orthognathic surgical planning and outcomes, as even minor variations can influence the surgical outcome [11,12].

Addressing these limitations, various machine learning methods have been proposed for jaw deformity diagnosis. These models, unlike traditional analyses reliant on predefined norms, aim to extract diagnostic patterns from training populations. Several studies have utilized Convolutional Neural Networks (CNNs) for diagnosis based on frontal/lateral cephalograms [13] or facial photographs [14] and also Multi-Layer Perceptrons (MLPs) using cephalometric measurements [15] or 3D coordinates of landmarks [16] as input for diagnosis. However, their reliance on extensive training data, compounded by variability in defining cephalometric norms, and the technical complexity requiring programming expertise, limit their accessibility to clinicians. Furthermore, the “black box” nature of these models, where interpretability is limited, worsens their utility in clinical settings.

Large Language Models (LLMs), propelled by advances in transformer architectures [17] and the vast internet-derived data [18], have become pivotal in artificial intelligence. Models like GPT-3 [19] demonstrate advanced text understanding and generation capabilities without task-specific fine-tuning. One remarkable feature of these models is their capacity for in-context learning (ICL) [20], allowing them to adapt to new tasks and data types, including diverse medical datasets [21]. The remarkable capabilities of LLMs have encouraged their application in the medical field [22], suggesting a potential revolution in improving diagnostic accuracy and clinical decision-making. This will potentially shift the paradigm of medical diagnosis, treatment planning, and patient interaction towards an integrated AI-assisted approach in healthcare [23].

This paper introduces a novel approach for diagnosing facial deformities using LLMs in cephalometric analysis, replacing the conventional training-test mechanism. By converting key measurements into descriptive texts and incorporating expert dental knowledge, the LLM provides grounded, adaptable, and intuitive outputs. This methodology advances facial deformity diagnosis research in three key areas:

### Educational Diagnostic Tool for Clinician Trainees

Our methodology aims to serve as a valuable educational tool for novice clinicians. By transforming complex cephalometric data into accessible text-based interpretations, the model provides not just data, but also contextual understanding of various diagnoses. This approach encourages trainees to compare and contrast their initial assessments with the model’s reasoning, exposing them to a broad spectrum of clinical scenarios. More importantly, it may help develop their ability to independently interpret complex data, fostering critical thinking and a more nuanced understanding of the complexities involved in diagnosing facial deformities as suggested in previous studies [24, 25].

### Enhancing Clinical Interpretability Accessibility

Clinical interpretability in our context refers to how clinicians can understand and apply the model’s outputs and underlying rationale. This feature is crucial as it provides clinicians, especially novices, with clear and relatable explanations that guide their diagnostic decisions. Additionally, our LLM-based methodology enhances accessibility by using natural language processing to interpret complex medical data through simple textual inputs with user-friendly web interfaces such as ChatGPT or Gemini, requiring only that clinician input patient-specific data into our pre-formulated prompt template. This eliminates the need for extensive setup or programming expertise, making it easily adaptable for various dental and medical fields suggested in [26], thus enhancing diagnostic precision and facilitating efficient decision-making.

### Feasibility and Future Potential

This study assesses the feasibility of using LLMs as diagnostic tools for complex medical data. While LLMs are not yet viable as standalone diagnostic tools, they are evolving rapidly. Innovations include the integration of multi-modality inputs and the expansion of training data, which are expected to significantly enhance model performance. Beyond these enhancements, a critical aspect that has emerged from prior research is the importance of prompt engineering. Our study contributes to this field by proposing structured prompts that can significantly enhance the utility of LLMs in clinical scenarios. These prompts are designed based on insights gathered from existing research and are tailored to improve the educational utility of models for trainee clinicians. Based on these findings, we propose structured prompts validated within our experimental settings. These prompts are intended to serve as a baseline for future models, potentially improving the efficacy of prompts in enhanced LLM architectures. Our approach aims to refine the utility of LLMs in clinical diagnostics, ensuring more precise and efficient outcomes for patient care.

## 2. Deformity Diagnosis with LLMs

### 2.1. Materials

In this in-silico study, we utilized anonymized retrospective data from Houston Methodist Hospital, focusing on patients with jaw deformities who had preoperative 3D models in AnatomicAligner® VSP software [27]. Patients with significant jaw asymmetry (mandibular midpoint over 3 mm or bilateral-point asymmetry over 4 mm) were excluded [28]. To improve the generalizability of our findings, we organized the data into temporal cohorts (Table 1). The initial cohort included patients who underwent surgery from August 2017 to April 2023, totaling 101 participants. The subsequent cohort comprised 50 patients who received surgery between May 2023 and March 2024. An experienced oral and maxillofacial surgeon evaluated the AnatomicAligner files to classify each patient’s mandibular position as normal, prognathic, or retrognathic, excluding any surgical simulation data for objectivity.

**Table 1.** Patient demographics of study cohorts.

|  | Initial cohort (N=101) |  |  |  | Subsequent cohort (N=50) |  |  |  | p-value |
| --- | --- | --- | --- | --- | --- | --- | --- | --- | --- |
|  | Retrognathic | Normal | Prognathic | All | Retrognathic | Normal | Prognathic | All |  |
| Age | 25.13±10.39 | 21.95±9.52 | 23.98±7.80 | 24.03±9.31 | 28.70±12.83 | 22.5±4.82 | 24.64±6.56 | 25.92±9.71 | 0.071<sup>1</sup> |
| Male | 9 | 12 | 21 | 42 (41.6%) | 4 | 7 | 8 | 19 (38.0%) | 0.806<sup>2</sup> |
| Female | 29 | 7 | 23 | 59 (58.4%) | 16 | 1 | 14 | 31 (62.0%) |  |
| SNB | 73.95±4.77 | 79.79±5.16 | 84.50±4.74 | 79.64±6.77 | 72.90±5.07 | 81.90±2.87 | 86.55±4.23 | 80.34±7.68 | 0.590<sup>3</sup> |
| FAng | 84.19±4.79 | 91.49±4.24 | 95.03±3.79 | 90.28±6.51 | 84.74±4.76 | 92.33±2.28 | 96.23±3.62 | 91.01±6.61 | 0.529<sup>3</sup> |
| MUL | 108.44±7.90 | 121.18±9.34 | 126.27±7.27 | 118.60±11.34 | 108.93±9.00 | 126.24±6.59 | 123.42±6.90 | 118.08±10.81 | 0.759<sup>1</sup> |

<sup>1</sup>Mann-Whitney, <sup>2</sup>Chi-square, and <sup>3</sup>Independent t-tests are conducted to compare between initial and subsequent cohort.

The Neutral Head Position (NHP) [29], a standard reference position in craniofacial assessments, was used to standardize each patient’s alignment. For LLM diagnosis, we selected three key cephalometric measurements linked to jaw deformity: SNB Angle, Facial Angle (FAng), and Mandibular Unit Length (MdUL) [16], as shown in Fig. 1.

**Fig 1.**
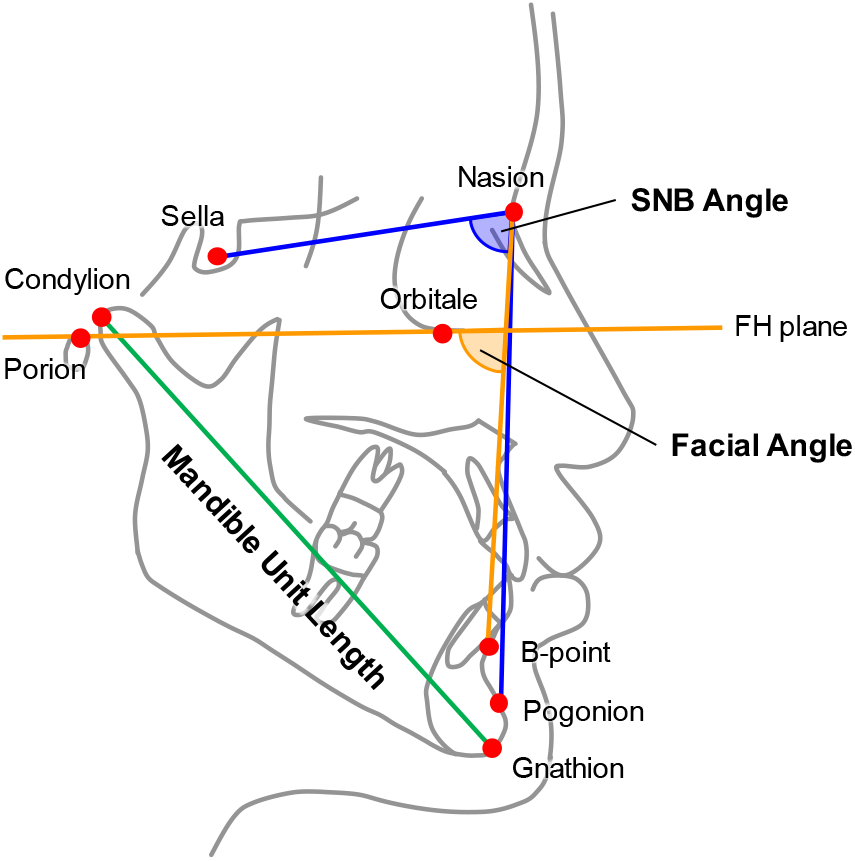
Cephalometric measurements: SNB Angle (Sella-Nasion to B-point Angle), Facial Angle (Angle between Nasion-Pogonion and the Frankfurt Horizontal Plane), and Mandible Unit Length (Distance between Condylion and Gnathion).

### 2.2. Prompt Engineering for Diagnosis

#### 2.2.1. Value Categorization to Text Description

LLMs excel in text generation due to training on extensive text corpuses but are less effective in directly processing numerical data like cephalometric measurements (e.g., SNB Angle, Facial Angle, MdUL) [30]. Therefore, we converted these numerical measurements into textual descriptions to align better with LLMs’ capabilities.

The conversion process involved:

1. Calculate the mean (*μ*) and standard deviation (*σ*) of the cephalometric measurement across normal subjects.
2. Defining adverbs (e.g., slightly, somewhat, moderately, considerably, extremely) to indicate the extent of deviation from the mean.
3. Categorizing measurement values based on their deviation from *μ* (in terms of *σ*) into specific textual intervals.

For example, an SNB Angle within [*μ*, *μ* + 0.5*σ*] can be expressed as “slightly above the mean of the normal subjects”, whereas a MdUL falls within [*μ* − 1.5*σ*, *μ* − 1.0*σ*] can be expressed as “moderately below the mean of the normal subjects”. Additionally, values outside the range of *μ* ± 2.0*σ* were considered extreme ranges. The range between *μ* − 2.0*σ* and *μ* + 2.0*σ* was uniformly divided into segments, varying from as few as 4 to as many as 14 segments. The division of cephalometric measurements values into discrete intervals can be found in Supplementary material I.

#### 2.2.2. Prompts for LLMs

The entire prompt for LLM is thoroughly presented in Supplementary material II. In the subsequent sections, we carefully examine each part of this prompt, providing insights into their distinct functions and importance in the field of jaw deformity diagnostics.

##### Role of LLM in Jaw Deformity Diagnostics

Advancements in LLMs have enhanced their capabilities in customization and role-playing, crucial for interactive and personalized tasks [31]. In line with these developments, we have configured our approach to specialize in the area of jaw deformity diagnostics. This specialization leverages the LLM’s enhanced interactive abilities to provide more accurate and context-specific diagnostic insights.

##### Definition of Anatomical Landmarks, Cephalometric Measurements, and Diagnostic Categories

To counter the variability in definitions due to LLMs’ diverse training data, our prompts include specific descriptions of anatomical landmarks, cephalometric measurements, and diagnostic categories. This ensures that LLM analyses align with standard practices in jaw deformity diagnostics.

##### Adverbs List from the Greatest to the Least Degree

The prompt uses a range of adverbs to label the extent of mandibular deviation, ordered from the most to the least degree: “*To describe the degree of mandibular deviation, the following adverbs are used from the greatest to the least degree: ‘extremely’, ‘considerably’, ‘moderately’, ‘somewhat’, ‘slightly’*.*”*

##### Goal of LLM in Deformity Diagnostics

While a lengthy prompt can contain a lot of information, it also runs the risk of weakening the focus on the final task to be performed by the LLM. To address this, by directly presenting the tasks that the LLM must perform, we can thereby steer the output towards precise, comprehensive, and clinically relevant results. *“I will provide you with the ranges for SNB, FAng, and MdUL. Your goal is to find the most suitable mandible category for the patient*.*”*

##### Important Guidelines for Mandibular Classification

In diagnosing jaw deformities, it’s essential to acknowledge the complex and detailed nature of the task. We utilize three pivotal cephalometric measurements for diagnosis, each with its own distinct level of importance and inherent characteristics. To ensure a precise and sophisticated analysis, the following cautionary guidelines are critical in our approach:

1. Avoid Simply Averaging Measurements: The LLM should not oversimplify the diagnostic process by averaging the values of these distinct measurements. Each measurement provides specific insights and contributes uniquely to the overall diagnosis. Averaging could lead to a loss of important diagnostic details.
2. Refrain from Majority Voting: The LLM must not use a majority voting for diagnosis. Such an approach might overlook the individual significance and contribution of each measurement, leading to an oversimplified and potentially inaccurate diagnosis.
3. Adhere to Singular Classification: Patients’ mandibles should be categorized into a single label, even in cases of conflicting measurements. The LLM should interpret these discrepancies instead of opting for a ‘mixed’ classification, ensuring that each diagnosis reflects a comprehensive understanding of the data.

By adhering to these guidelines, the LLM is directed to engage in a more comprehensive, detailed analysis, reflecting the depth and complexity of traditional diagnostic processes. This ensures that the LLM-based diagnosis remains aligned with the sophisticated clinical decision-making inherent.

##### Few-Shot Learning Examples for Enhanced LLM Performance

In-context learning with LLMs demonstrates strong few-shot performance across various NLP tasks including input-label pairing and example selection strategies [32]. In the context of our study, we leverage this few-shot learning approach by providing specific measurement information for each patient, paired with their corresponding mandible label. This strategy enables the LLM to more accurately identify and categorize jaw deformities with a relevant set of examples.

##### Mandible Categories (PROGNATHIC, NORMAL, RETROGNATHIC)

In our diagnostic approach, we explicitly define the categories of mandible conditions we aim to diagnose. By clearly outlining these categories, we guide the LLM to narrow down its analysis and identify the most appropriate category for each case. This method plays a crucial role in streamlining the diagnostic process, as it allows the LLM to focus its computational capabilities on distinguishing between these predefined categories.

##### Answer Formatting

In our diagnostic approach using LLMs, we utilize chain-of-thought reasoning [33] to effectively process complex tasks [30]. The LLM begins by analyzing each cephalometric measurement individually, ensuring a thorough understanding of each data point. It then integrates these individual analyses to form a comprehensive view, allowing for a nuanced evaluation that takes all factors into account. Finally, the LLM synthesizes this information to classify the diagnosis, ensuring that the final determination is both precise and contextually informed. Answer formatting is crucial for facilitating these logical steps, structuring the process to enable the LLM to deliver informed and reliable diagnostic conclusions.

## 3. Experiments and results

### 3.1 Implementation and Configuration

In this study, we utilized multiple LLMs to evaluate our approach for diagnosing jaw deformities, including:

- Three variants of the open source LLAMA-2 models by Meta AI, namely “LLAMA-2-7B-Chat”, “LLAMA-2-13B-Chat”, and “LLAMA-2-70B-Chat” [34]. These models were selected for their diverse capacities in text interpretation.
- Two GPT models by OpenAI, “gpt-3.5-turbo-1106” and “gpt-4-1106-preview” [35]. These GPT models are known for their advanced text processing and generation capabilities.
- The “Gemini-Pro” [36] model by Google DeepMind was also utilized, known for its proficiency in handling complex language processing tasks.

The training process was confined to the initial cohort, with the data divided into training and test sets. The subsequent cohort was strictly used for testing to assess the generalizability and effectiveness of the models across temporal variations, ensuring a robust evaluation of the performance.

We employed random sampling for few-shot learning across 10 different random seeds to reduce variability. This produced 10 prediction sets per seed, aggregated for a comprehensive performance analysis. Moreover, we maintained the ratio between different classes to ensure stable performance from an in-context learning perspective.

For deterministic outcomes, we set the LLMs’ ‘top_p’ parameters to 0.0. This adjustment guarantees deterministic results by exclusively selecting the words with the highest probability. Experiments with closed-source LLMs (GPT models, Gemini model) were conducted via their APIs, while open-source LLAMA-2 models were run on six NVIDIA A100 (40GB) GPUs. Responses from LLMs for all patients are presented in Supplementary Material III.

### 3.2 Evaluating LLMs Against Various Diagnostic Approaches

Our study compared the performance of Threshold-based, Machine Learning models, and LLMs in diagnosing jaw deformities. Threshold-based methods utilized recalibrated cut-off values for SNB, Facial Angle, and MdUL tailored to our patient cohort [37]. For machine learning, we used models such as SVM, Random Forest, XGBoost, and MLP, training them on the same 30-patient dataset as the LLMs, with 20% of the data for validation.

In LLM experimentation, we set the number of discrete intervals to eight, as depicted in the segment labeled ‘8C’ in Supplementary material I, and used 30 few-shot learning examples, based on our ablation study findings. Performance was measured by balanced accuracy (the average of accuracy on each class) and F1-score (macro), considering class imbalance.

Results (Table 2) showed that the Facial Angle (FAng) threshold method achieved the highest balanced accuracy in the initial cohort with a balanced accuracy of 65.26±2.23 and an F1-score of 61.12±1.87. In the subsequent cohort, the SNB threshold method led the way, achieving the top balanced accuracy of 68.55±3.23 and an F1-score of 63.38±6.27. Among the machine learning models, MLP showed and RandomForest showed the best performance in the initial and subsequent cohort, respectively. In the LLM category, LLAMA-2-13B model demonstrated strong and consistent performance across both cohorts. It achieved a balanced accuracy of 62.52±5.01 and an F1-score of 62.46±3.93 in the initial cohort, and significantly improved in the subsequent cohort to a balanced accuracy of 67.68±8.15 and an F1-score of 67.46±7.31. GPT-4 model also showed consistent performance, with balanced accuracies of 62.69±3.12 in the initial cohort and 63.15±7.80 in the subsequent cohort, and F1-scores of 62.80±3.04 and 62.56±7.95, respectively. Notably, there were no significant performance differences across cohorts for most LLM and Machine Learning Models, with exceptions noted in threshold-based methods.

**Table 2.**
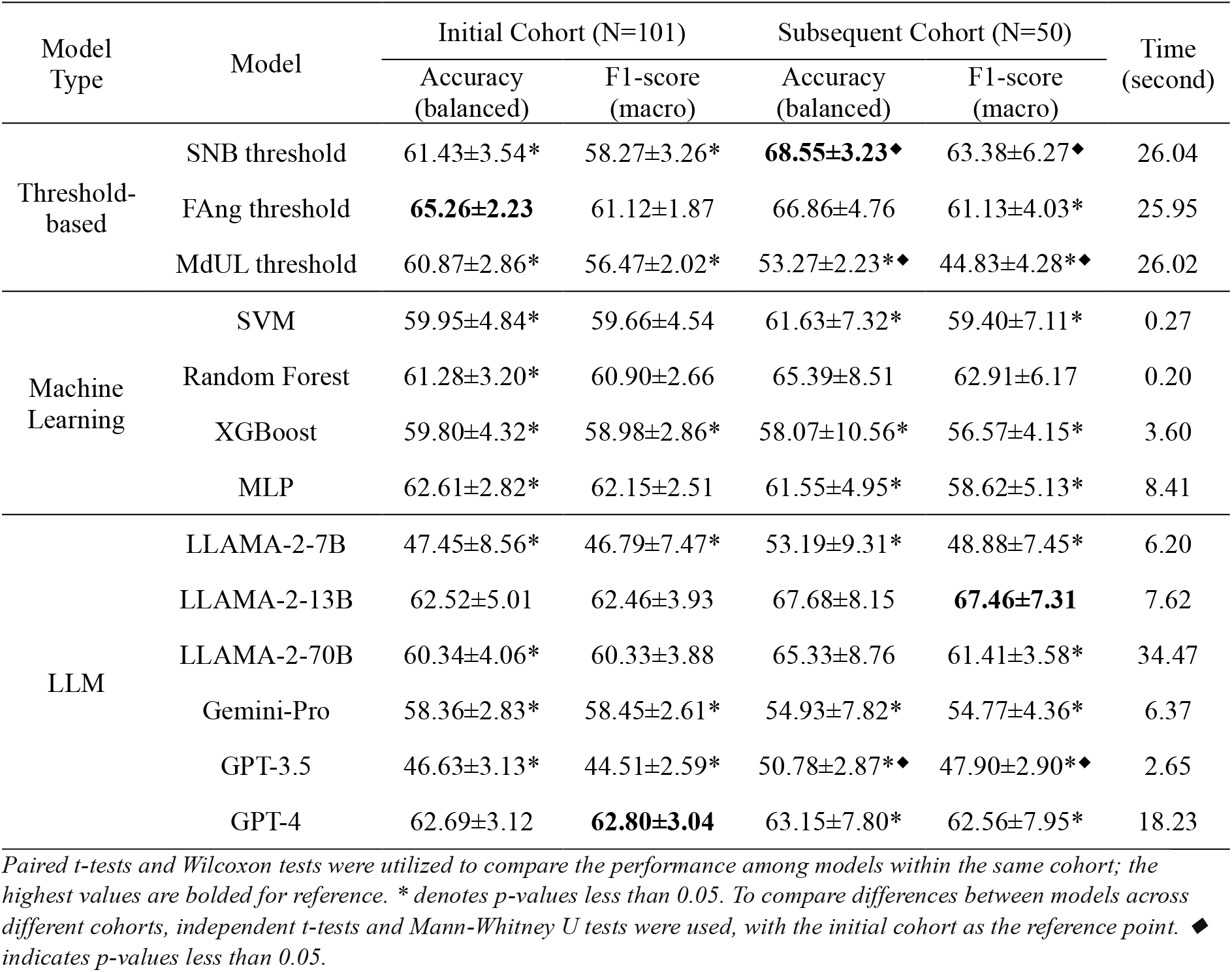
Comparative performance of Threshold-based methods, Machine Learning models, and LLMs in cephalometric diagnosis.

Additionally, we have measured the time taken by each model. For the Threshold-based method and Machine Learning models, we recorded the time required for training (excluding the time for hyperparameter search). Since LLMs do not require a traditional training phase, we focused on measuring the inference time per patient for these models. Specifically, this refers to the average time the model takes to diagnose one patient, based on input that includes 30 few-shot learning examples and the measurements of the patient in question. The larger LLM models took a comparable amount of time to the Threshold methods whereas the smaller LLM models required less than 10 seconds, similar to Machine Learning models (XGBoost, MLP). To ensure uniform experimental conditions, all experiments were performed on a server equipped with eight A100 GPUs and an AMD EPYC 7502 32-Core CPU.

### 3.3 Optimizing LLM Performance: An Ablation Study

#### 3.3.1. Impact of the Number of Segments

The ablation study assessed how different segment numbers in the categorization process (from numerical to text format, as in section 2.2.1) affected F1-scores (Fig 2). In this experiment, we applied the LLAMA-2-7B and LLAMA-2-13B models. For the smaller LLAMA-2-7B model, we observed no significant performance difference across different number of segments. However, with the larger LLAMA-2-13B model, there was a noticeable difference in performance as the number of few-shot learning examples increased, especially between groups with fewer segments (4C, 6C) and those with more (8C, 10C, 12C, 14C). Based on these findings, and considering a balance between efficiency and performance, we selected 8C as the optimal number of segments for our experiment.

**Fig 2.**
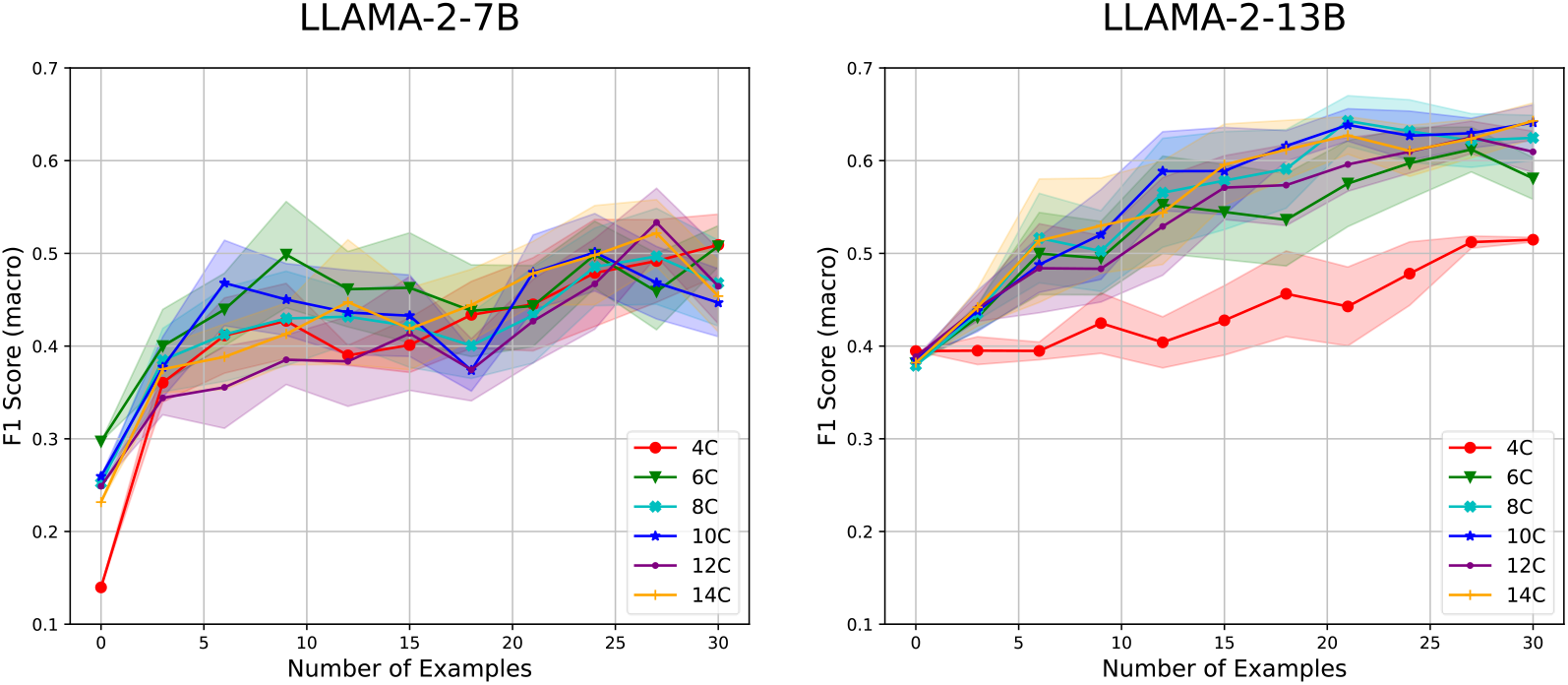
Performance across varying number of segments to determine optimal selection.

#### 3.3.2. LLM Comparison and Impact of the Number of Examples

In our study, we solely rely on the inference phase of LLMs to achieve diagnostic outcomes. When only target patient’s cephalometric measurements are provided as input, the LLM bases its diagnosis on the foundational knowledge it has acquired, likely from dental literature or publications. However, this does not guarantee that the information reflects consistent population characteristics, necessitating a shift in the data’s information context. By including multiple examples of input features alongside their corresponding classes from our dataset with target patient’s cephalometric measurements, we anticipate the LLM to adapt to our specific dataset through this process.

To identify the optimal number of examples required for this adaptation, we conducted an ablation study where we varied the number of examples from 0 to 30 in increments of 3 and observed the performance changes. In this experiment, we expanded our investigation to include additional LLMs: LLAMA-2-70B, GPT-3.5, GPT-4, and Gemini-Pro, comparing their performance (Fig 3). In zero-shot learning scenario, larger models like GPT-4, LLAMA-2-70B, and Gemini-Pro outperformed smaller ones. Performance improved with more few-shot learning examples, particularly in smaller models like LLAMA-2-7B and 13B, highlighting their sensitivity to additional contextual information. All models tended to converge in performance effectiveness with around 30 examples, leading us to determine that 30 examples optimize performance across different LLMs. While increasing this number could further enhance performance, the limitation of input token size makes it impossible to compare all models, hence we set 30 as the maximum of this experiment.

**Fig 3.**
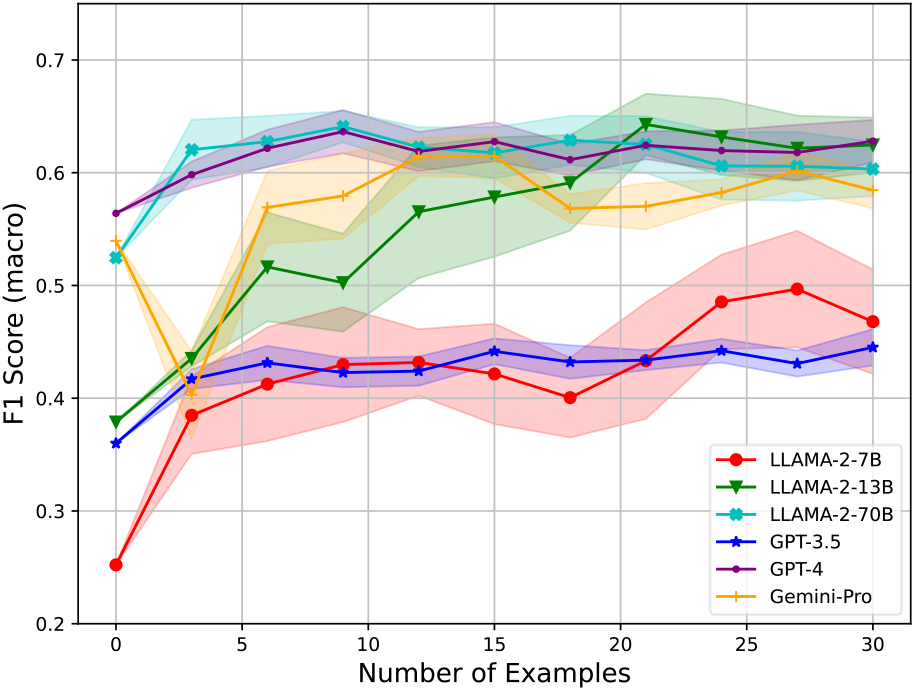
Impact of few-shot learning examples on performance across different LLMs.

#### 3.3.3. Comparative Analysis of Input Types in LLMs: Text Descriptions Versus Numerical Values

Our study hypothesized that LLMs would interpret text descriptions more effectively than direct numerical values. To verify this, we compared the performance of these two input types under a fixed setting derived from our prior ablation study: the number of segments equal to 8 and a set of 30 examples. For a fair comparison, numerical inputs included the mean (*μ*) and standard deviation (*σ*) of normal subjects. Excluding the smallest model, LLAMA-2-7B, in all other cases, the use of text description as input demonstrated higher performance in both balanced accuracy and F1-score compared to the use of numerical values as input (Table 3).

**Table 3.** Performance comparison of LLMs using text description and numerical values as inputs.

| Input Type | Model | Accuracy (balanced) | F1-score (macro) |
| --- | --- | --- | --- |
| Numerical Value | LLAMA-2-7B | 52.59±5.20* | 46.90±2.78* |
|  | LLAMA-2-13B | 42.95±5.89* | 40.19±4.58* |
|  | LLAMA-2-70B | 51.30±3.48* | 49.94±3.99* |
|  | Gemini-Pro | 58.20±3.12* | 57.71±2.88* |
|  | GPT-3.5 | 44.74±3.25* | 43.07±2.40* |
|  | GPT-4 | 60.08±2.45* | 59.87±2.35* |
| Text Description | LLAMA-2-7B | 47.45±8.56* | 46.79±7.47* |
|  | LLAMA-2-13B | 62.52±5.01 | 62.46±3.93 |
|  | LLAMA-2-70B | 60.34±4.06 | 60.33±3.88 |
|  | Gemini-Pro | 58.36±2.83* | 58.45±2.61* |
|  | GPT-3.5 | 46.63±3.13* | 44.51±2.59* |
|  | GPT-4 | **62.69±3.12** | **62.80±3.04** |

Paired *t*-tests and Wilcoxon tests compare results; the highest values are bolded as reference points. \* denotes *p*-values less than 0.05.

### 3.4 Proportion of Ambiguous Classification

While LLM outputs offer flexible sentence-formatted responses, we encountered an issue with ambiguous classifications that didn’t fit our predefined categories, such as “MIXED mandible” or “PROGNATHIC-RETROGNATHIC mandible”. To address this, we analyzed the proportion of ambiguous responses across all LLMs used (Fig 4).

**Fig 4.**
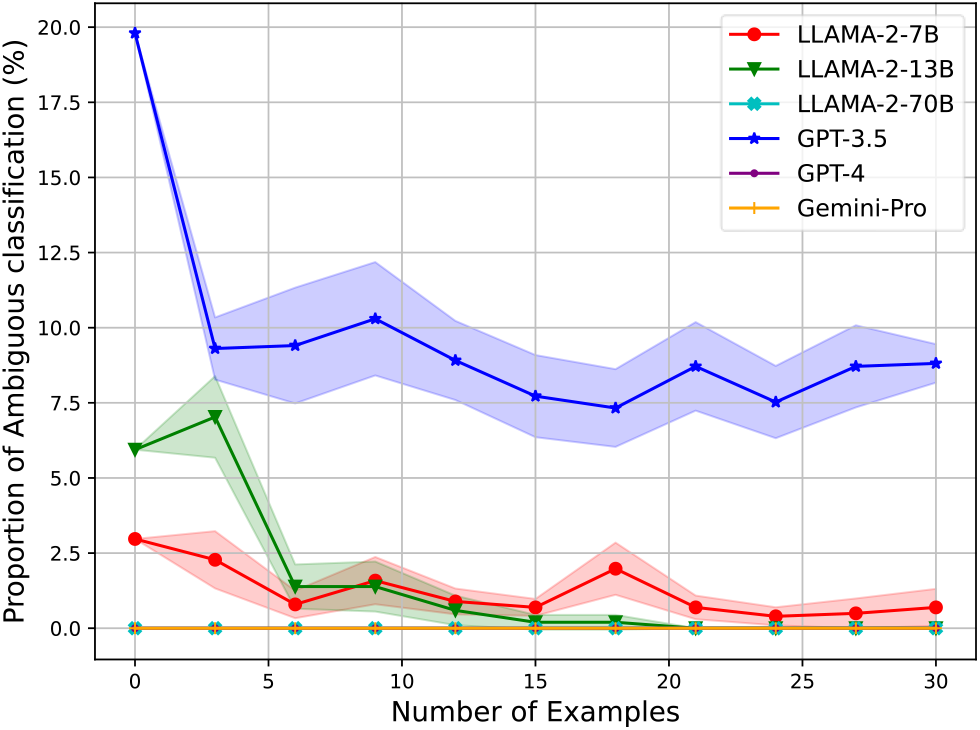
Analysis of ambiguous classification proportion in various LLMs with increasing example inputs.

**Fig 5.**
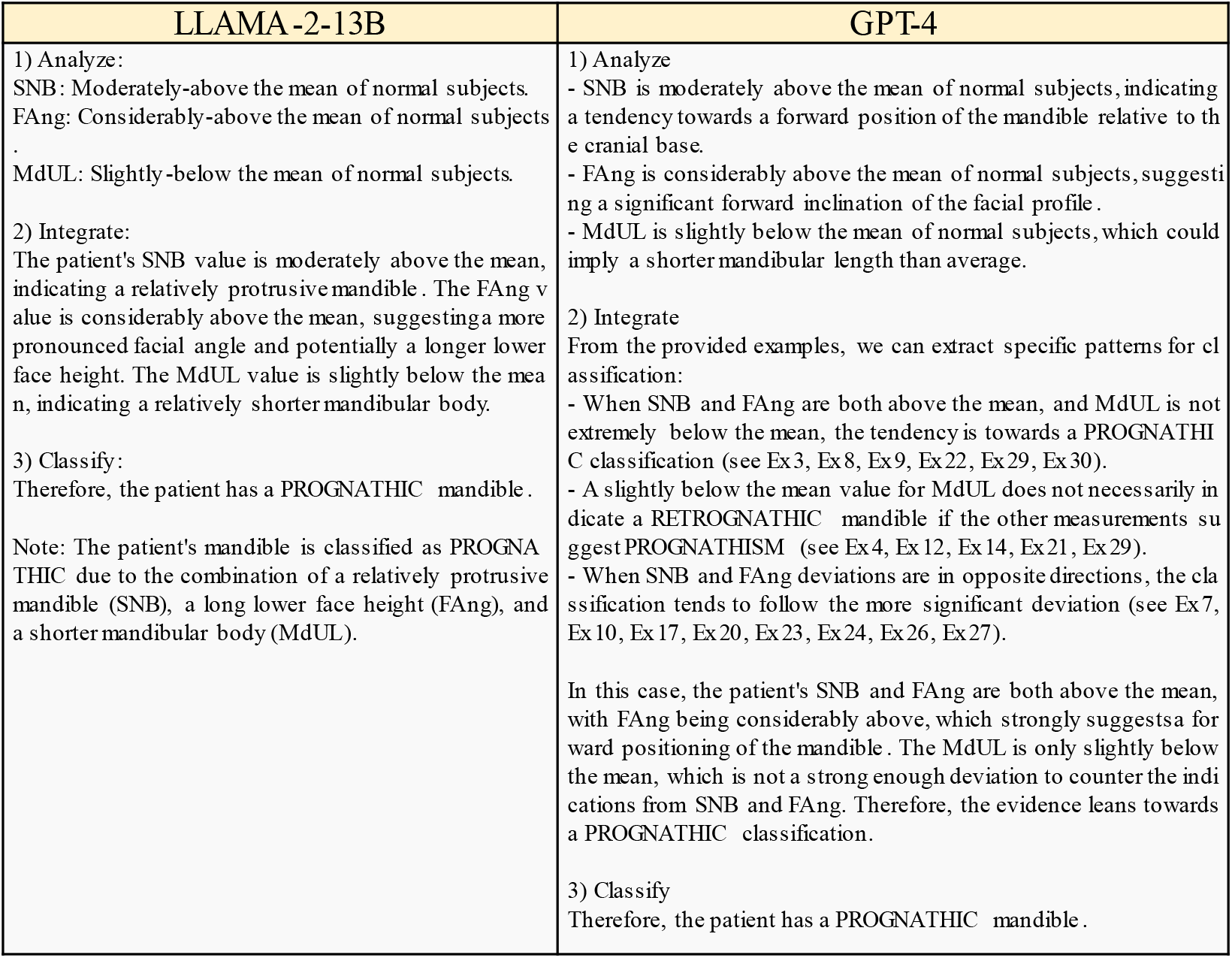
Responses of LLMs for an example case. Responses for all patients can be found in supplementary material III.

We found that larger LLMs (LLAMA-2-70B, GPT-4, Gemini-Pro) did not produce ambiguous classifications. In contrast, smaller models showed varying levels of ambiguous responses, which decreased as more examples were provided. Notably, LLAMA-2-13B’s ambiguous classifications dropped from 6% to 0% with 21 examples, indicating a relationship between the number of examples and the clarity of classification.

## 4. Discussion

Our study explores the potential of using LLMs for diagnosing jaw deformities, with a particular focus on the performance of larger models in zero-shot and few-shot learning scenarios. These models demonstrate rapid performance saturation with fewer examples in few-shot learning, indicating a robust pre-existing knowledge base that supports diagnostic tasks. This finding suggests that LLMs are well-suited to adapting to specific requirements in medical diagnostics.

One of the pivotal achievements of our study is the conversion of cephalometric measurements into text format, converting measurement values into intuitive adverbial descriptions, aligning the raw data with the strengths of natural language processing. Our method standardizes the interpretation of these measurements into a text-based format, reducing potential misinterpretations and simplifying the diagnostic process; crucial for ensuring that the approach is accessible and adaptable to various clinical scenarios.

In evaluating the best model for clinical implementation, we prioritized criteria such as generalizability and clinical interpretability, where GPT-4 exceeded others. Our findings highlight that, besides their robust performance, the most significant contribution of LLMs lies in their ability to enhance the training experience of novice clinicians. This is achieved by providing clear, context-rich interpretations and rationales for various diagnoses, thereby aiding in the establishment of diagnostic guidelines.

Furthermore, the clinical interpretability of our methodology stands out as a considerable advantage. We emphasize that our model, while not currently suitable as a standalone diagnostic tool, serves as an invaluable educational aid. It helps clinician trainees understand and apply the reasoning behind diagnostic conclusions, offering a diverse set of perspectives that enrich their learning experience.

Additionally, the text-based nature of LLMs enhances their accessibility, particularly for clinicians who may not be well-versed in complex programming or machine learning systems. This accessibility facilitates the integration of these advanced diagnostic tools into routine clinical practice, making them especially valuable for less experienced clinicians.

While this LLM-based approach is proposed as an educational tool for less experienced clinicians, a more thorough validation is required from multiple angles to verify its rationality thoroughly. Furthermore, the potential issues arising from an over-reliance on LLM information must be critically addressed. These directions necessitate essential follow-up research to ensure the responsible and effective use of LLMs in clinical settings.

As we continue to refine our approach, recognizing that the current accuracy level may not be suitable for standalone clinical decision-making is crucial. The insights gained from analyzing a limited set of cephalometric measurements indicate that expanding the dataset to include a broader range of diagnostic inputs could enhance performance. This aligns with findings from a recent study [16], which employed 50 landmark 3D coordinates in an MLP model to achieve higher accuracy, demonstrating the benefits of incorporating more comprehensive input data. Moreover, the reliance on statistical norms derived from specific databases may introduce biases that could affect the accuracy and representativeness of the outcomes. Continuous refinement and adjustment are necessary to ensure the model accurately reflects diverse patient groups.

Finally, considering that the LLMs used were general-purpose models trained on broad datasets, there is potential for developing domain specific LLMs. These specialized models could offer improved diagnostic accuracy in medical fields such as dentistry, representing a promising direction for future research.

## 5. Conclusion

In conclusion, our study demonstrates the potential of using LLMs to diagnose jaw deformities, particularly highlighting the effectiveness of larger models in zero-shot and few-shot learning scenarios. By converting cephalometric measurements into intuitive text formats, we have significantly enhanced the accessibility and clinical interpretability of diagnostic processes, which is especially beneficial for less experienced clinicians. Although our LLMs do not yet meet the accuracy required for standalone clinical decision-making, they serve as valuable auxiliary tools that improve the understanding and application of diagnostic data. Moving forward, refining these models to incorporate broader and more specifically targeted datasets, and adapting them to leverage domain-specific medical data, could significantly improve their accuracy and utility in clinical settings, potentially transforming the landscape of medical diagnostics.

## Supporting information

Supplementary Material III

## Data Availability

Not applicable.

## Acknowledgements

This work was partially supported by the National Institutes of Health under award R01DE021863.

## Declarations

### Ethical Approval

The study was approved by the Institutional Review Board of Houston Methodist Hospital (IRB # Pro00013802).

