## Supplementary Material I for "Large Language Models Diagnose Facial Deformity"

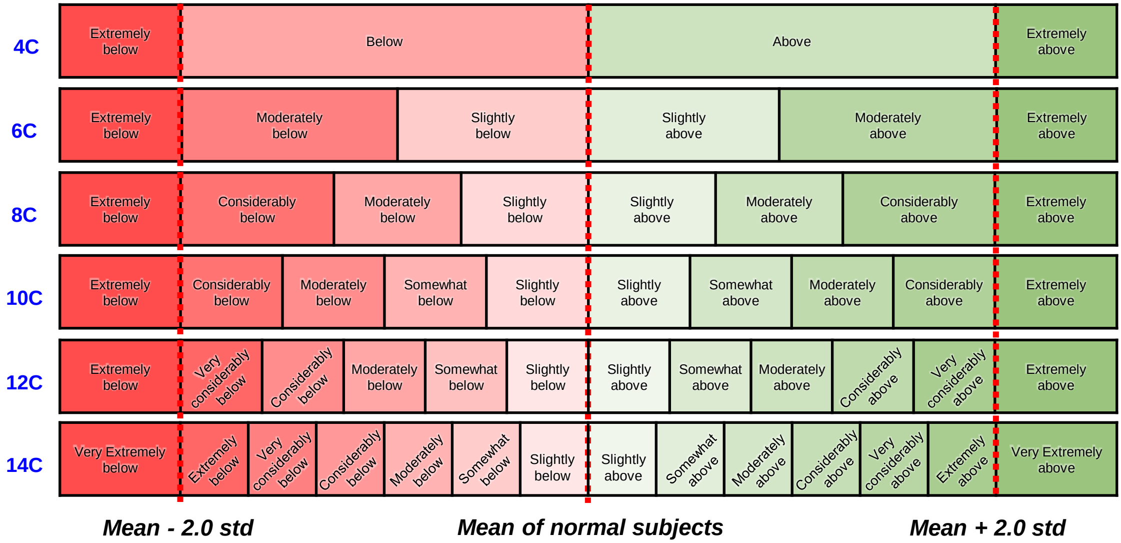


**Supplementary Material I** Division of cephalometric measurements values into discrete intervals based on the mean and standard deviation of normal subjects.
